# Genetic risk score-informed re-evaluation of spirometry quality control to maximise power in epidemiological studies of lung function

**DOI:** 10.1101/2024.07.31.24311269

**Authors:** Jing Chen, Nick Shrine, Abril G Izquierdo, Anna Guyatt, Henry Völzke, Stephanie London, Ian P Hall, Frank Dudbridge, SpiroMeta Consortium, CHARGE Consortium, Louise V Wain, Martin D Tobin, Catherine John

**Author notes:** Joint corresponding authors (Jing Chen), (Catherine John).

## Abstract

**Background and aim:** Epidemiological studies of lung function may discard one-third to one-half of participants due to spirometry measures deemed “low quality” using criteria adapted from clinical practice. We aimed to define new spirometry quality control (QC) criteria that optimise the signal-to-noise ratio in epidemiological studies of lung function.

**Material and methods:** We proposed a genetic risk score (GRS) informed strategy to categorize spirometer blows according to quality criteria. We constructed three GRSs comprised of SNPs associated with forced expiratory volume in 1 second (FEV_1_), forced vital capacity (FVC) and the ratio of FEV_1_ to FVC (FEV_1_/FVC) in individuals from non-UK Biobank cohorts included in prior genome-wide association studies (GWAS). In the UK Biobank, we applied a step-wise testing of the GRS association across groups of spirometry blows stratified by acceptability flags to rank the blow quality. To reassess the QC criteria, we compared the genetic association results between analyses including different acceptability flags and applying different repeatability thresholds for spirometry measurements to determine the trade-off between sample size and measurement error.

**Results:** We found that including blows previously excluded for cough, hesitation, excessive time to peak flow, or inadequate terminal plateau, and applying a repeatability threshold of 250ml, would maximise the statistical power for GWAS and retain acceptable precision in the UK Biobank. This approach allowed the inclusion of 29% more participants compared to the strictest ATS/ERS guidelines.

**Conclusion:** Our findings demonstrate the utility of GRS-informed QC to maximise the power of epidemiological studies for lung function traits.

## Introduction

Impairment of lung function, as measured by spirometry, is central to chronic respiratory diseases including chronic obstructive pulmonary disease (COPD), but also predicts mortality in the general population[1]. Genome-wide association studies (GWAS) have proven to be an effective tool for identifying genetic variants that are associated with complex diseases and traits, providing valuable insights into disease biology and informing development of diagnostic tools and potential treatments[2]. For lung function, the most recent GWAS identified associated genetic variants explaining 33% of the heritability of FEV_1_/FVC (less for FEV_1_ and FVC)[3], indicating that additional associations could be found in more powerful studies.

However, GWAS of lung function may discard one-third to one-half of participants due to spirometry measures deemed “low quality”, substantially limiting the potential sample size[4]. Spirometry measures the flow and volume of air over time, typically in a forced expiratory manoeuvre, i.e. a vigorous, complete exhalation following maximal inhalation. **Figure 1** illustrates key spirometric parameters. Spirometry is effort and technique-dependent, and thorough quality control (QC) is considered essential to ensure measurements are unaffected by inadequate technique or artefacts (such as a cough). However, it has been suggested that “Pulmonary function standards are not static. They should be questioned. There is always room for improvement in any set of pulmonary function standards”[5, 6]. Moreover, QC for epidemiological studies may not require the same level of stringency as clinical practice, where results are used for diagnosis and management of individual patients. Hankinson *et al* . have previously suggested visual inspection of blow curves by human reviewers in addition to computer assessment to avoid unnecessary rejection of valid data[7]. While this approach could enhance the inclusion of valid tests, it may become impractical for large-scale studies and could be subject to bias.

**Figure 1:**
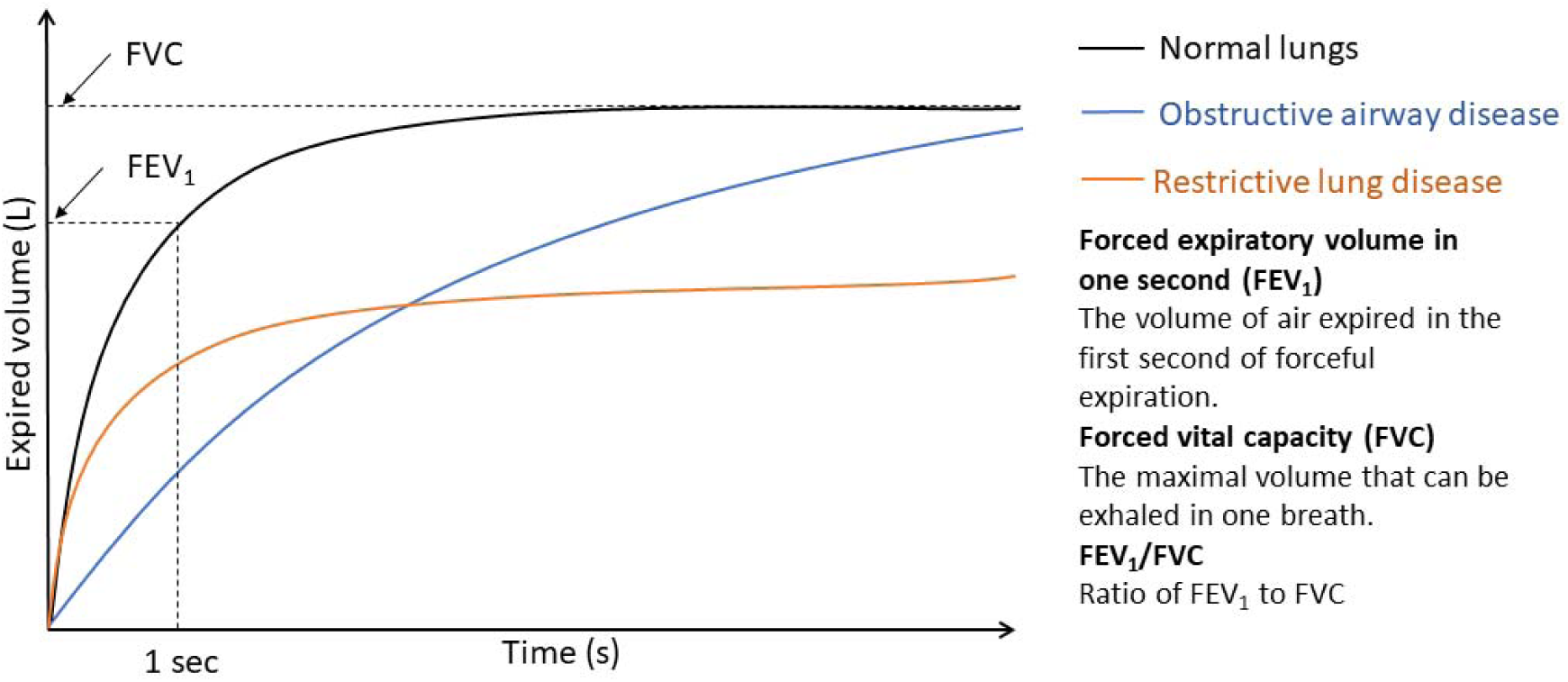
Volume-time curve. The curve plots the total volume of air expired by time, from full inspiration until full expiration. Normal blow shows rapid increase in volume of air expired initially, then curve forms a plateau; Obstructive blow shows prolonged increase but ends the same point; Restrictive blow shows rapid increase as normal, but curve forms a plateau much sooner.

Genetic information can predict individual continuous traits such as lung function by tallying the number of risk alleles for each individual to give a genetic risk score (GRS) (sometimes called polygenic risk score, PRS, or polygenic score, PGS, **Figure 2**) [8]. These may include hundreds to millions of genetic variants and are typically weighted by the size of association of each allele with the trait of interest[8]. By checking concordance between the predicted lung function values based on genetics and the actual measured lung function traits, we can reassess the value of spirometer blows that were previously deemed as “low quality”. In this study, we aimed to establish new spirometry QC criteria informed by GRS to optimise the signal-to-noise ratio in epidemiological studies.

**Figure 2:**
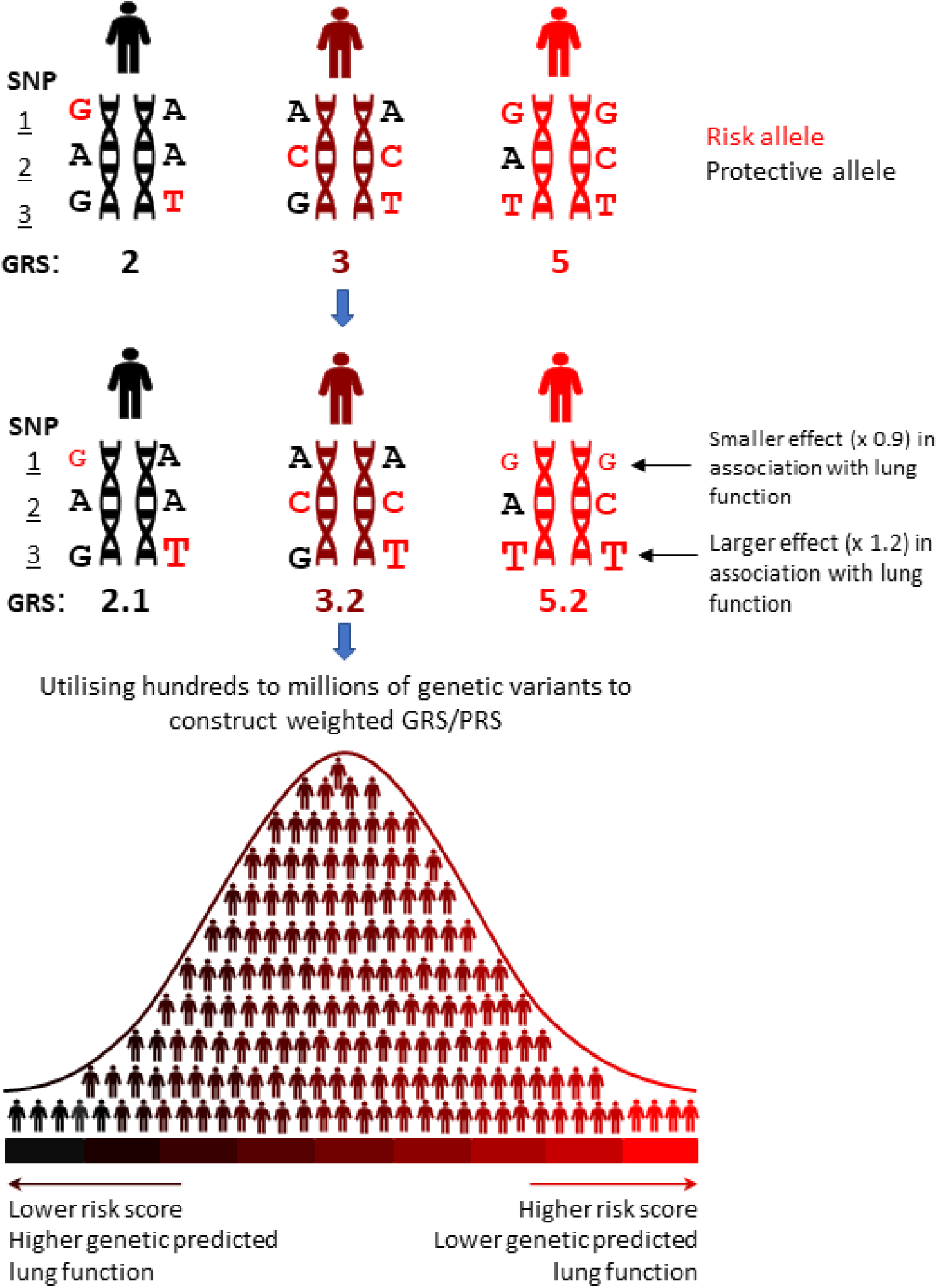
Basic principle of constructing GRS/PRS. Schematic showing the principles of constructing a GRS. Top section (a) shows a theoretical example of an unweighted risk score is calculated at three SNPs, assuming an additive model; middle section (b) shows a weighted risk score for the same alleles, whereby the score for each allele is weighted according to its association with lung function; bottom section (c) shows a normal distribution of individual scores for a GRS/PRS using hundreds to millions of genetic variants, where the score at each variant is weighted according to its association with the trait of interest (lung function), ideally in an independent population.

## Methods

### Spirometry quality control

We undertook analyses in the UK Biobank European population [9]. In UK Biobank, each individual was asked to perform up to three blows on a Vitalograph spirometer (Vitalograph Pneumotrac 6800). These blows were used to derive measures of lung function, including the forced expiratory volume in 1 second (FEV_1_), forced vital capacity (FVC) and the ratio of FEV_1_ to FVC (FEV_1_/FVC), from a spirogram (volume-time curve) recorded by the Vitalograph software.

A blow received an automated error code from the spirometry software if: (1) there was hesitation (excessive extrapolated volume[10] at the start of the blow, coded “START”); (2) the time to peak flow was excessive (“EXPFLOW”); (3) a cough was detected during the manoeuvre (“COUGH”); (4) there was not an adequate plateau at the end of the blow (“END”) (see **Table 1** for additional detail). It was also possible for the spirometer operator to explicitly reject the blow (“REJECT”). In previous GWAS of lung function, blows were deemed unacceptable and excluded from analysis if they had any of the above error codes[4]. Additionally, as in previous GWAS[3, 4], we checked each blow for inappropriate negative values which indicate a problem with the blow (for any of: forced expiratory flow at 25% of FVC; forced expiratory flow at 75% of FVC; average forced expiratory flow between 25% and 75% of the FVC; peak flow; extrapolated volume; volume at forced expiratory time) (coded “NEGATIVE”), the start of the blow underwent a further check for hesitation (“START2”), and consistency (<5% difference) was checked between the FEV_1_ and FVC output from the spirometer and that rederived from the spirogram (“CONSISTENCY”). In this study, we identified blows that would previously have failed QC due to any of the above error codes or additional QC steps, and labelled these with corresponding “acceptability flags” (**Table 1**).

**Table 1:** Reasons for spirometer blows failing quality control .

| **2019**<br>**ATS/ERS**<br>**criterion** | **Definition of**<br>**ATS/ERS**<br>**criterion (2019)** | **Acceptability**<br>**flags in this**<br>**study** | **Acceptability flag definition in this**<br>**study** | **Number of**<br>**individuals**<br>**with blows**<br>**failing within**<br>**acceptability**<br>**flags stratum** |
| --- | --- | --- | --- | --- |
| Acceptable<br>start of<br>blow (i.e.<br>an<br>immediate,<br>explosive<br>start with<br>no<br>hesitation) | Extrapolated<br>volume <5% or<br>100ml (150ml in<br>2005 criteria) | START<br>(spirometer<br>output) | Present if the extrapolated volume<br>at start of test is excessive | 54,922 |
|  |  | START2<br>(additional<br>QC) | Present if the extrapolated volume is<br>not less than 5% of FVC or 150 ml,<br>whichever is greater. | 53,840 |
|  |  | EXPFLOW<br>(spirometer<br>output) | Present if time to peak flow is<br>excessive | 164,319 |
| No cough<br>in first<br>second (for<br>FEV<sub>1</sub>) | N/A | COUGH<br>(spirometer<br>output) | Present if a cough was detected<br>during the manoeuvre | 11,857 |
| Acceptable<br>end of<br>blow | Adequate<br>terminal plateau<br>(<25ml flow in<br>last 1s) or<br>length of blow<br>>=15s or FVC<br>meets<br>repeatability<br>threshold | END<br>(spirometer<br>output) | Present if an adequate plateau at the<br>end of the test does not exist | 103,092 |
| N/A | N/A | REJECT<br>(spirometer<br>output) | Present if the spirometer operator<br>explicitly rejects the blow | 1,610 |
| N/A | N/A | NEGATIVE<br>(additional<br>QC) | Present if inappropriate negative<br>values derived from the blow for any<br>of: forced expiratory flow at 25% of<br>FVC; forced expiratory flow at 75%<br>of FVC; average forced expiratory<br>flow between 25% and 75% of the<br>FVC; peak flow; extrapolated<br>volume; volume at forced expiratory<br>time. | 577 |
| N/A | N/A | CONSISTENCY<br>(additional<br>QC) | Present if the difference between<br>the FEV<sub>1</sub> and FVC output from the<br>spirometer and that rederived from<br>the spirogram was greater than 5%. | 3,183 |

In previous lung function GWAS and in our baseline analysis for this study, a repeatability threshold of 250ml was applied.

### Testing the association between GRSs and lung function traits derived from spirometer blow measurements

We calculated the GRSs for European-ancestry individuals in UK Biobank using European-specific weights trained from Shrine *et al* . [3] (**Supplementary Methods**). In the UK Biobank European population, we then conducted iterative testing of the GRS association within groups of spirometry blows stratified by acceptability flags, to identify and rank the flags most likely to cause failure of spirometry blow quality (**Figure 3**). For each lung function trait, we first stratified all the blows by acceptability flag (**Figure 3, step I, Supplementary Methods**). Where lung function measures within a stratum showed no significant association with the GRS (P>0.05), we considered that the corresponding acceptability flag to indicate unacceptable blow quality and thus a reason for exclusion. For the remaining blows, we applied an iterative selection process to identify the acceptability flags that were most likely to cause failure of spirometry blow quality as shown in **Figure 3 (Step II)**. Based on the outcome of this, we ranked the remaining acceptability flags according to their impact on the spirometer blows in our new approach (see Results for details).

**Figure 3:**
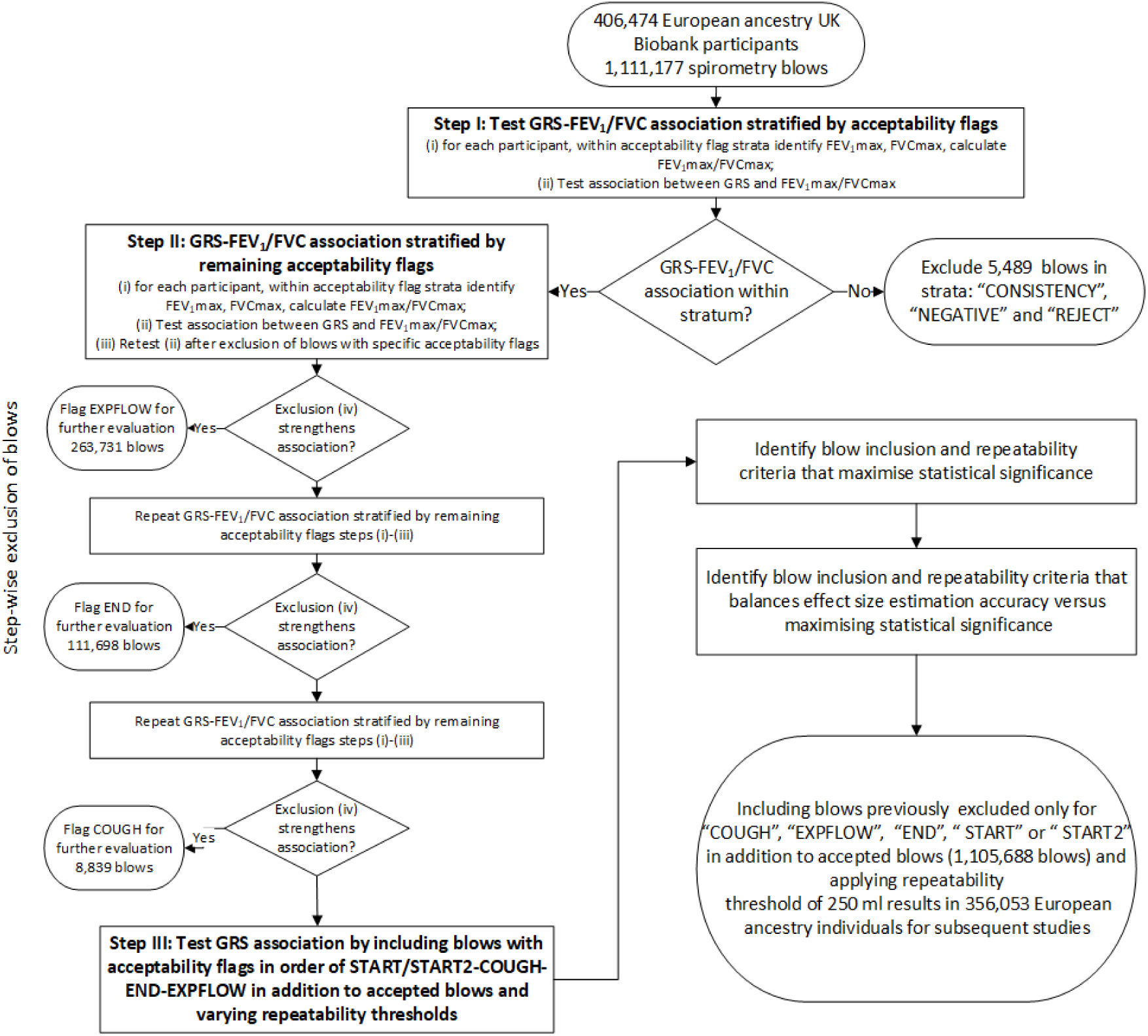
Flowchart for evaluating spirometer blow quality and spirometry QC criteria. Definitions of acceptability flags (“COUGH”, “EXPFLOW”, “END”, “START”, “START2”, “CONSISTENCY”, “NEGATIVE”, “REJECT”) were given in Table 1.

### Re-evaluating spirometry QC criteria for association study

Based on the grouping of blows above, we then aimed to identify which acceptability and repeatability criteria would maximise the association of the lung function measures with GRS as measured by the level of statistical significance (Z-score for the association) (**Figure 3, Step I**)**I**.**I** We varied our acceptability criteria by including blows with acceptability flags in order of the ranking generated in the previous step, in addition to blows previously considered acceptable. We also varied our repeatability threshold (from 150ml to 400ml in 50ml increments). Repeatability was based on the difference from any other blow, even if that blow was not accepted. Using these different criteria, we tested the association with the GRS.

To ensure that effect size estimates retained acceptable precision whilst maximising statistical significance, we then tested genetic associations with the sentinel SNPs known to be associated with lung function[3], using the different acceptability criteria and repeatability thresholds described above, and examined the effect sizes and P-values for sentinels of lung function signals using these different QC criteria. All lung function traits were untransformed, adjusting for age, age^2^, height, smoking status, and relatedness (mixed models in BOLT-LMM[11]).

### Assessing the credibility of GWAS findings

To illustrate the gain in power using our newly defined QC criteria in epidemiological studies, we conducted a GWAS in the UK Biobank European population as an example to compare GWAS findings with those found using the previous QC criteria. Credibility of newly identified signals was assessed using a Bayesian framework previously described by Okbay *et al* [12]and Turley *et al* [13] (**Supplementary methods**).

## Results

In UK Biobank, 445,541 individuals had at least two measures of FEV_1_ and FVC, along with complete information for age, sex, standing height and derived smoking status (smoking status derived by Shrine *et al* . 2019[4]. Of these individuals, 406,474 were assigned as European ancestry using k-means clustering and ADMIXTURE v1.3.0[14] as described in Shrine *et al*[4]. We used FEV_1_ and FVC measurements (UK Biobank Field IDs 3063 and 3062), along with the blow curve time series measurements (UK Biobank Field ID 20031) and the Vitalograph spirometer blow quality metrics (UK Biobank Field ID 20031).

### Evaluating spirometer blows quality with different acceptability flags

Using the previous, more stringent approach to spirometry QC, we identified 713,885 spirometer blows as “accepted” for 354,746 European individuals. The best measures of FEV_1_/FVC derived from “accepted” blows for these individuals were strongly associated with the GRS (β_SD_change_of_GRS_=0.246, 95% CI [0.243, 0.249], P<1.00E-300) (**Figure 4a**).

**Figure 4:**
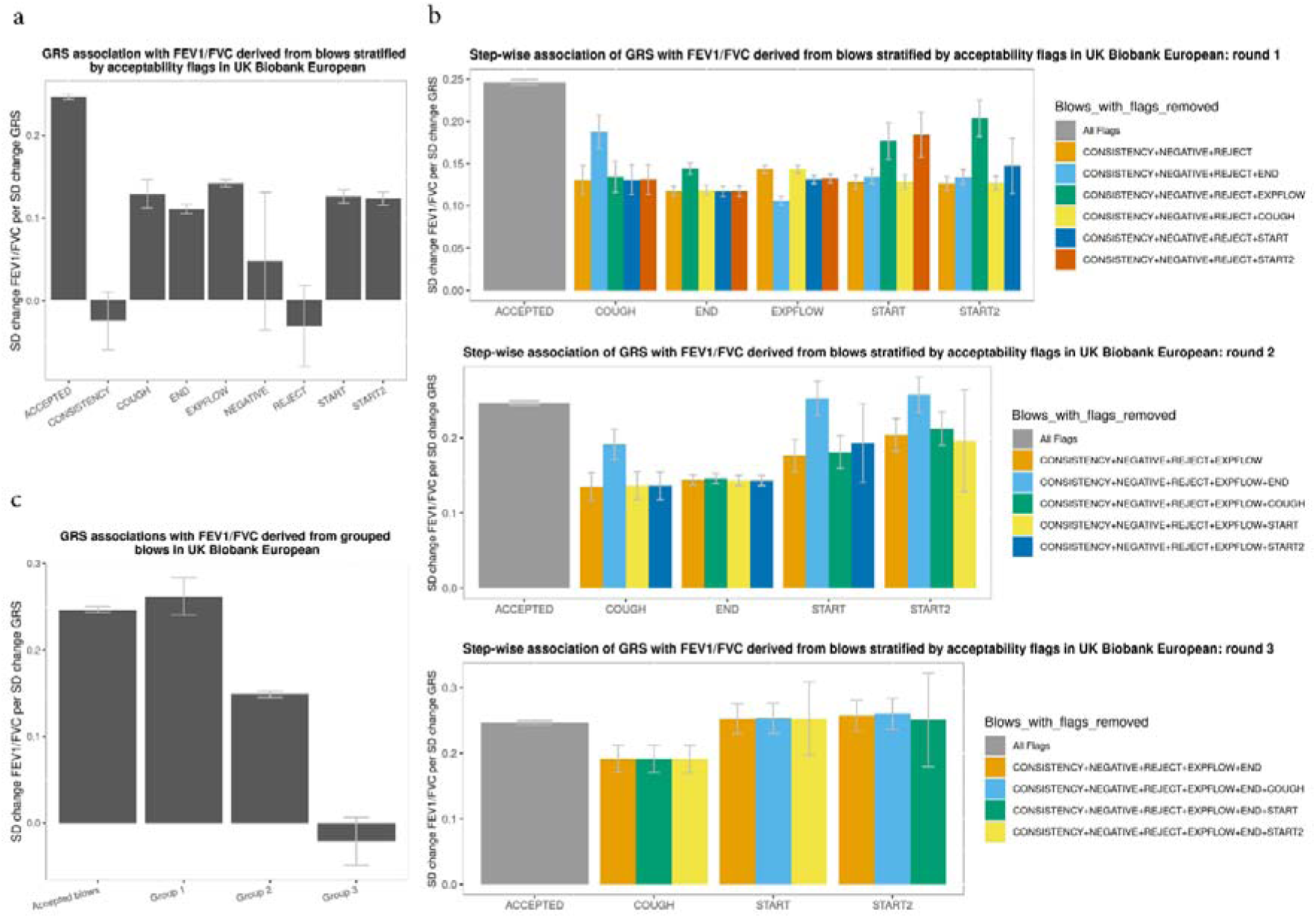
Ranking the impact of acceptability flags on spirometer blows. **a,** GRS association with FEV_1_/FVC derived from spirometer blows stratified by acceptability flags shown as the s.d. change in FEV_1_/FVC per s.d. increase in GRS. N.B. A blow could be included in multiple acceptability flag strata if it carries multiple acceptability flags. **b**, Iterative selection process. **c**, GRS association with FEV_1_/FVC derived from grouped spirometer blows shown as the s.d. change in FEV_1_/FVC per s.d. increase in GRS. Group 1 represents blows with hesitation flags only (“START” or “START2”). Group 2 represents blows with cough, end-of-blow or time to peak flow flags (“COUGH”, “END” or “EXPFLOW”) but without flags of “REJECT”, “CONSISTENCY” or “NEGATIVE”. Group 3 represents blows with acceptability flags of “REJECT”, “CONSISTENCY” or “NEGATIVE”. The height of the bars shows the point estimate of the effect and whiskers show the 95% CI.

From the association of GRS with the FEV_1_/FVC derived from blows in each of our acceptability flag strata, blows with acceptability flags “CONSISTENCY”, “NEGATIVE” or “REJECT” were not associated with the GRS (**Figure 4a**), indicating that these acceptability flags represent unacceptable spirometry blow quality (β_SD_change_of_GRS_=-0.022, 95% CI [-0.049, 0.006], p=0.1292, **Figure 4c, Group 3**). To rank the remaining acceptability flags that cause failure of the spirometry blow quality in descending order of severity, we conducted an iterative selection process (**Figure 4b**). In the first round of selection (round 1), we identified excessive time to peak flow (“EXPFLOW”) as the next most likely cause of failure of spirometry quality. This was based on the observation that additionally removing blows with “EXPFLOW” flag led to an increase in the magnitude of the effect size estimate in the association results in all the remaining strata. Similarly, we identified an inadequate terminal plateau (“END”, round 2) and cough (“COUGH”, round 3) as the next most likely to cause failure of spirometry quality in the subsequent rounds of selection. Collectively, blows from groups “EXPFLOW”, “END” and “COUGH” were associated with GRS but with a smaller effect size than previously accepted blows (β_SD_change_of_GRS_=0.149, 95% CI [0.145, 0.153], p<1.00E-300, **Figure 4c, Group 2**), after removing blows with the flags excluded in previous steps. For the remaining flags relating to hesitation (“START” and “START2”), the magnitude of the effect size of GRS association in the corresponding strata (β_SD_change_of_GRS_=0.262, 95% CI [0.240, 0.284], P=1.14E-118, **Figure 4c, Group 1**) was similar to that from the acceptable blows (β_SD_change_of_GRS_=0.246, 95% CI [0.243, 0.249], P<1.00E-300, **Figure 4c, Accepted blows**), after removing blows with all the other acceptability flags.

For FEV_1_ and FVC, we observed similar results to those for FEV_1_/FVC above in ranking the spirometer blow quality but with the “COUGH” flag showing a similar effect size to accepted blows (**Supplementary Figure 1 and Supplementary Figure 2**). For FVC, “END” had a larger impact than “EXPFLOW”. Since the results suggest acceptability flags have greater impact on the measurements of FEV1/FVC than FEV1 and FVC, we based our subsequent analyses on FEV_1_/FVC.

### Re-evaluating spirometry QC criteria for association studies

We investigated the trade-off between sample size and measurement error by re-evaluating various inclusion criteria for spirometry QC. To do this, we included blows with varying acceptability flags with their inclusion ordered according to the findings above, and applied different repeatability thresholds to assess their impact on the association between GRS and FEV_1_/FVC. We found that including blows previously excluded for cough (COUGH), hesitation (START/START2), excessive time to peak flow (EXPFLOW) or lack of terminal plateau (END) in addition to accepted blows and applying a repeatability threshold of 350ml reached the maximum statistical significance in the association between GRS and FEV_1_/FVC. However, as expected, the effect size in the association results attenuated toward zero as QC criteria were successively relaxed (**Table 2**).

**Table 2:** The GRS association with FEV _1_/FVC derived from blows with varying inclusion criteria and repeatability threshold.

| **Blow inclusion criteria** | **Repeatability threshold(ml)** | **Beta\*** | **LCI (95% CI)** | **UCI (95% CI)** | **z score** | **Sample size N** |
| --- | --- | --- | --- | --- | --- | --- |
| No acceptability flags | 250 | 0.266 | 0.263 | 0.269 | 162.99 | 320591 |
| No acceptability flags OR hesitation acceptability flags only ("START" or "START2") | 150 | 0.270 | 0.267 | 0.274 | 154.54 | 275957 |
|  | 200 | 0.268 | 0.264 | 0.271 | 160.67 | 304931 |
|  | 250 | 0.266 | 0.263 | 0.270 | 164.06 | 321724 |
|  | 300 | 0.264 | 0.261 | 0.267 | 164.82 | 331685 |
|  | 350 | 0.262 | 0.259 | 0.265 | 164.85 | 337893 |
|  | 400 | 0.260 | 0.257 | 0.263 | 164.70 | 342155 |
| No acceptability flags OR hesitation or cough acceptability flags only ("START", "START2", "COUGH") | 150 | 0.270 | 0.267 | 0.274 | 154.69 | 276696 |
|  | 200 | 0.268 | 0.264 | 0.271 | 160.82 | 305834 |
|  | 250 | 0.266 | 0.263 | 0.269 | 164.25 | 322734 |
|  | 300 | 0.264 | 0.261 | 0.267 | 165.00 | 332806 |
|  | 350 | 0.262 | 0.259 | 0.265 | 165.12 | 339076 |
|  | 400 | 0.260 | 0.257 | 0.263 | 165.00 | 343379 |
| No acceptability flags OR hesitation, cough or end-of-blow acceptability flags only ("START", "START2", "COUGH", "END") | 150 | 0.265 | 0.262 | 0.269 | 154.03 | 285804 |
|  | 200 | 0.263 | 0.259 | 0.266 | 160.14 | 317024 |
|  | 250 | 0.261 | 0.258 | 0.264 | 163.56 | 335310 |
|  | 300 | 0.258 | 0.255 | 0.262 | 164.28 | 346314 |
|  | 350 | 0.256 | 0.253 | 0.259 | 164.36 | 353359 |
|  | 400 | 0.255 | 0.252 | 0.258 | 164.36 | 358243 |
| No acceptability flags OR hesitation, cough, end-of-blow or time to peak flow acceptability flags only ("START", "START2", "COUGH", "END", "EXPFLOW") | 150 | 0.265 | 0.262 | 0.268 | 157.47 | 299668 |
|  | 200 | 0.262 | 0.258 | 0.265 | 163.95 | 334952 |
|  | 250 | 0.260 | 0.257 | 0.263 | 167.59 | 356163 |
|  | 300 | 0.257 | 0.254 | 0.260 | 168.43 | 369039 |
|  | 350 | 0.255 | 0.252 | 0.258 | 168.69 | 377568 |
|  | 400 | 0.253 | 0.250 | 0.255 | 168.49 | 383545 |

\* stands for standard deviation (SD) change of FEV<sub>1</sub>/FVC per SD change of GRS

To balance accuracy of effect size estimation versus maximising statistical significance (z-score) for genome-wide association studies, we examined the changes in the effect sizes and P values of the sentinels of FEV_1_/FVC signals identified by Shrine *et al*[3]. We found that additionally including blows previously excluded for cough (COUGH), hesitation (START/START2), excessive time to peak flow (EXPFLOW) or lack of terminal plateau (END), and applying a repeatability threshold of 250 ml optimized the signal-to-noise ratio in genetic association testing (**Figure 5**). Based on this finding, we proposed a new spirometry QC strategy for epidemiological studies, which retained 29% more participants using strictest ATS/ERS guidelines and increased the sample size from 275,084 to 356,053 individuals.

**Figure 5:**
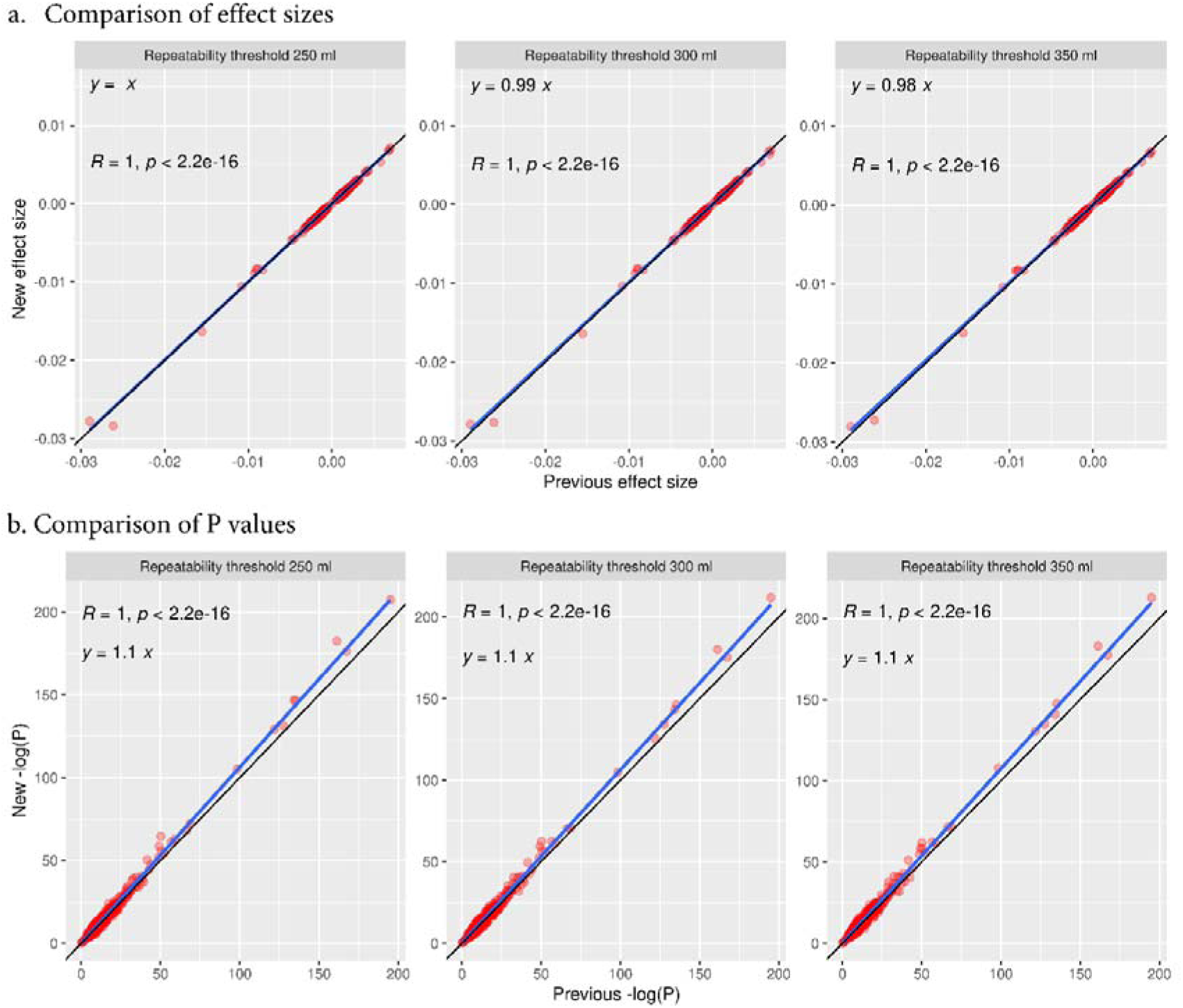
Examine the genetic association results of FEV _1_/FVC signals estimated from relaxed blow inclusive criteria at varying repeatability threshold of 250 ml, 300 ml and 350 ml. **a,** compare the effect sizes of FEV_1_/FVC signals using relaxed spirometry QC (New effect size, including blows previously only excluded for cough, hesitation, excessive time to peak flow or lack of terminal plateau (“START”, “START2”, “COUGH”, “END”, “EXPFLOW”) in addition to accepted blows) to the result obtained from previous spirometry QC (Previous effect size, only including accepted blows). **b**, compare the p values of FEV_1_/FVC signals using relaxed spirometry QC (New -log(P)) to the result obtained from previous spirometry QC (Previous -log(P)).

### Illustrate the gain in power using newly defined QC criteria

Using the newly defined spirometry QC criteria, the UK Biobank sample size increased from 320,591 in the most recent GWAS of lung function to 356,053, an 11% gain in sample size, and the mean χ^2^statistic (i.e. squared z scores for SNP and FEV_1_/FVC association) from GWAS increased from 1.27 to 1.29, leading to 15 additional sentinel SNPs associated with FEV_1_/FVC (selected 2 Mb regions centred on the most significant variant for all regions containing a variant with P <5×10^-9^, additional compared with analysis of UK Biobank alone using previous QC criteria[3]) (**Supplementary Table 2**). Of these, 7 were not identified in the largest GWAS of lung function to date. Examining the nearest genes to the sentinel SNPs, we found two novel genes in addition to 13 genes reported either in the most recent GWAS[3] or the EMBL-EBI GWAS Catalog. One of the novel genes, *FPR3*, encodes the formyl peptide receptor 3, a paralog of formyl peptide receptor 1. The functional role of *FPR3* is not fully understood. However, it is expressed in a range of immune cells, including macrophages and eosinophils, but not neutrophils, so has been hypothesized to play a role in allergic disease[15], and has also been associated with asthma and white blood cell counts in GWAS. We also found that enrichment of a previously implicated pathway[3], ESC pluripotency pathway, was strengthened by the newly identified gene *WNT16*.

For the newly identified hits, we followed procedures previously described by Okbay *et al.* [12] to calculate the posterior probability of true association, which exceeded 99% for all 15 additional loci for any assumption about the prior probability of non-null SNPs in the range 1% to 99%. To test the sentinel SNPs for replication, we used meta-analysis results from 42 European ancestry cohorts (excluding participants from UK Biobank) of 249,114 individuals generated by Shrine *et al*[3]. Due to the limited statistical power for replication of genome-wide significant association with a smaller sample size, we applied methods to assess the replication of the effect size of sentinel SNPs. For the set of 13 newly identified sentinel SNPs for FEV_1_/FVC available in the meta-analysis results, we regressed the effect sizes in UK Biobank on the effect sizes in the meta-analysis results with intercept constrained to be zero, after correcting the UK Biobank effect size estimates for winner’s curse bias using the method described in Turley *et al* [13]. The regression slope was 0.75 (standard error = 0.1376), being statistically significantly greater than zero (one-sided P=1.474×10^-4^) but not statistically distinguishable from one (one-sided P=0.0943), suggesting that the newly identified sentinel SNPs were replicated in independent datasets.

## Discussion

QC of spirometric measurements involves a range of metrics and criteria which are designed to ensure that clinical decision-making is based on accurate and reproducible measurements. Spirometry is also widely used in epidemiological research, including genetic association studies. Leading experts in spirometry and its QC have previously noted that, “it is unclear when quality is insufficient for acceptance of results into research studies”, and in particular have noted that end-of-test criteria may be applied too stringently[7]. The solution proposed of visual inspection of spirometry curves is not feasible in large-scale association studies, and in this context unnecessary exclusions may impact on power for novel discovery. We propose a new GRS-based method to define a QC strategy which maximizes power whilst maintaining acceptable precision, enabling an 29% increase in sample size using strictest ATS/ERS guidelines in UK Biobank. This identified 15 additional genetic loci not found in analysis of UK Biobank using the previous QC criteria, of which 7 were not identified in the largest GWAS of lung function to date, and two implicated novel genes highlighting new biology of interest. Eight were already identified in the largest consortium GWAS of lung function to date [3], but demonstrate that these discoveries could have been made earlier.

In this study, we introduce an iterative selection process to rank acceptability flags in descending order of their impact on spirometer blow quality failure. We then included blows with varying acceptability flags and repeatability threshold to refine the spirometry QC criteria for GWAS, with the aim to optimize the signal-to-noise ratio. Through the application of this strategy to lung function traits in UK Biobank, we demonstrated that the statistical power of GWAS can be increased by employing more inclusive spirometry QC criteria, as evidenced by substantial improvements in sample size, mean chi-square statistics and the identification of additional genetic association signals. These signals implicated two novel genes for lung function, one of which (*FPR3*) plays a role in innate immunity and has been implicated in asthma and allergic disease.

To date, although 1020 genetics signals have been discovered for lung function traits, much of the genetic contribution to lung function remains unexplained[3]. The problem of “missing heritability” could explained by undetected common variant associations and rare variant associations. The approach we outline here will boost power for GWAS of common/ rare variants such as those now available through the whole genome sequencing of UK Biobank [16], structural genomic variants as long read sequencing data become available. While the QC criteria established in UK Biobank may not be directly transferable to other studies, the same methodology can be applied. Our approach will be especially relevant where sample sizes are limited, such as in under-represented ancestries in genomic studies[17]. Furthermore, whilst genetic data are used to inform the spirometry QC, the increase in sample size and power from our approach is could be applicable to a wide range of epidemiological research questions, such as assessing lung function associations with environmental factors or biomarker levels. Biomarker measures are becoming more available as biotechnology evolves. Moreover, this methodology could potentially be applicable to other complex traits requiring intricate QC steps.

One limitation of our study is that we were not able to assess all ATS/ERS spirometry QC criteria. To fully meet 2019 ATS/ERS QC criteria, blows must also be free from glottic closure and from evidence of technical issues (faulty zero-flow setting, leak or obstruction). These are not generally included in automated spirometer output. Additionally, the analysis could benefit from a more powerful PRS, which would be expected to yield superior prediction performance.

In summary, our study highlights a useful application of GRS in epidemiological studies of lung function. GRS-informed QC boosts sample size and power for epidemiological studies, as illustrated by our discovery of new genetic associations for lung function. R scripts implementing this analysis are available from https://github.com/legenepi/GRS_informed_QC.

## Data Availability

All data produced in the present study are available upon reasonable request to the authors

https://github.com/legenepi/GRS_informed_QC

## Supplementary Data

### Supplementary Methods

#### Constructing GRS and testing its association with lung function traits

We built European-specific GRSs in a multi-ancestry study for FEV_1_, FVC and FEV_1_/FVC consisting of 425, 372 and 442 autosomal signals (genome-wide significant threshold of P < 5 × 10^-9^) associated with each trait respectively, using weights estimated from a multi-ancestry meta-regression in 244,472 individuals across 42 cohorts (**Supplementary Table 1**) that are independent from the UK Biobank[3]. We calculated the GRSs for 406,474 European-ancestry individuals in UK Biobank The associations between GRSs and lung function traits were tested using a linear model, adjusted for age, age squared, sex, height, smoking status and ten principal components.

For the first step of GRS association with lung function traits stratified all the blows by acceptability flag, a blow could be included in multiple strata if it had multiple acceptability flags. Within each acceptability flag stratum, we identified the best lung function measures (i.e. the best measure was defined as the highest measure for FEV_1_ and FVC, whilst FEV_1_/FVC was derived from the selected FEV_1_ and FVC) for each participant and tested the association of the GRS with the measured lung function trait to identify which strata were significantly associated with the GRS.

#### Bayesian framework to assess the credibility of GWAS findings

For genome-wide association testing under an additive model using BOLT-LMM[11], we used untransformed residuals from linear regression of lung function traits against age, age^2^, sex, height, smoking status. To assess the credibility of our findings from the genetic association study using the relaxed spirometry QC criteria, we used a standard Bayesian framework previously described by Okbay *et al*[12] and Turley *et al*[13]. Briefly, we used maximum likelihood to fit the SNP effect from the GWAS result using a mixture of a Gaussian prior with a point mass at zero. The prior for effect size corresponding to any given SNP *j* is

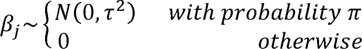

where *τ*^2^ is the prior belief of the variance of effect size of non-null SNPs and *π* is the prior belief of the fraction of non-null SNPs. For each assumed prior *π* from 1% to 99%, we used the maximum likelihood estimates of parameters to calculate the posterior probability that a SNP is non-null given its estimated effect size and given that it is significant. To assess the replication of the effect size in independent study, the effect size for each SNP is corrected for winner’s curse by

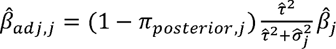

where *π_posterior,j_* is the posterior probability of the SNP *j* being non-null and 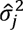 is the squared standard error of the GWAS estimates for SNP *j*.

### Supplementary Figures

**Supplementary Figure 1:**
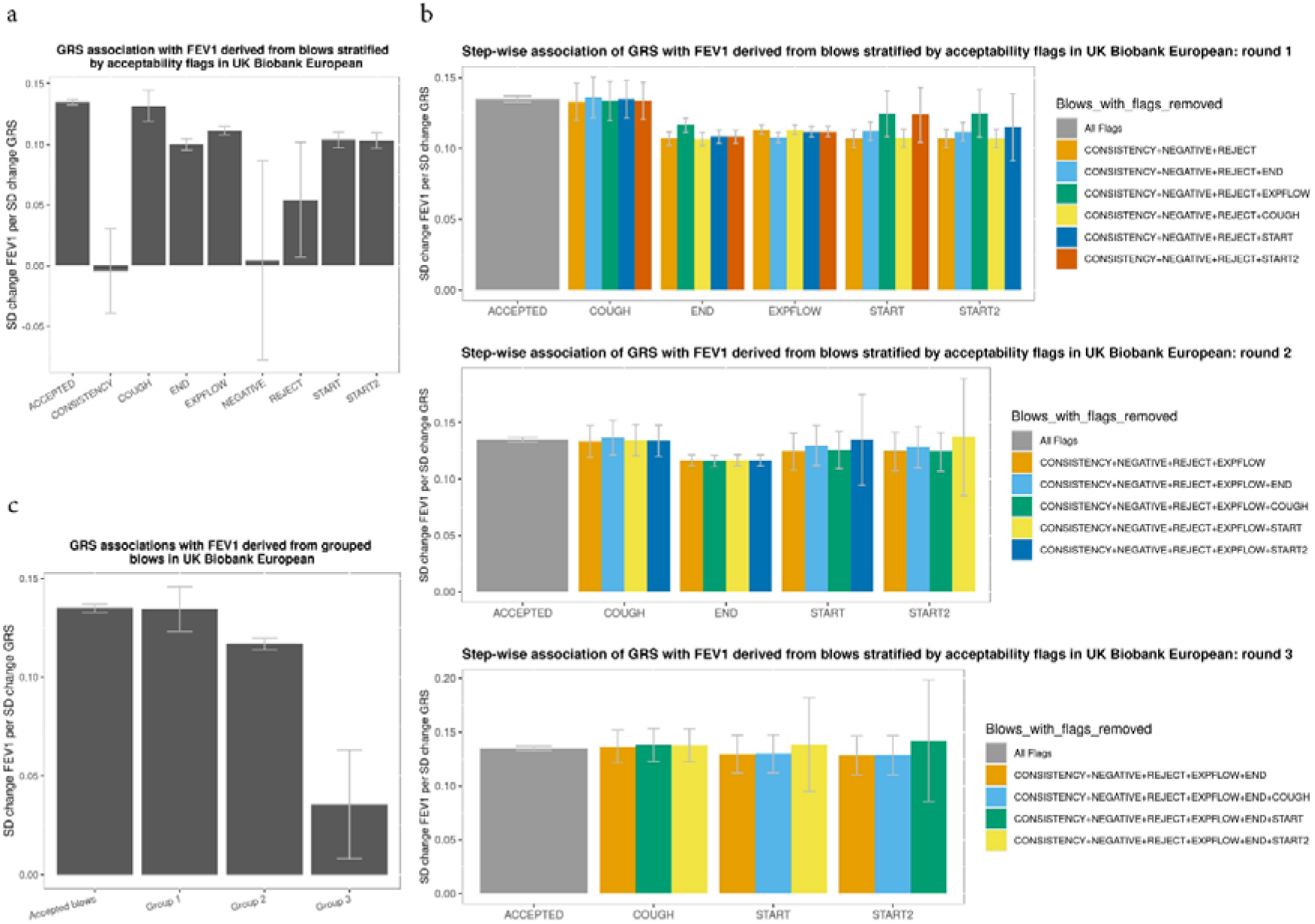
Ranking the impact of acceptability flags on spirometer blows. **a,** GRS association with FEV_1_ derived from spirometer blows stratified by acceptability flags shown as the s.d. change in FEV_1_ per s.d. increase in GRS. N.B. A blow could be included in multiple acceptability flag strata if it carries multiple acceptability flags. **b**, Iterative selection process. **c**, GRS association with FEV_1_ derived from grouped spirometer blows shown as the s.d. change in FEV_1_ per s.d. increase in GRS. Group 1 represents blows with hesitation flags only (“START” or “START2”). Group 2 represents blows with cough, end-of-blow or time to peak flow flags (“COUGH”, “END” or “EXPFLOW”) but without flags of “REJECT”, “CONSISTENCY” or “NEGATIVE”. Group 3 represents blows with acceptability flags of “REJECT”, “CONSISTENCY” or “NEGATIVE”. The height of the bars shows the point estimate of the effect and whiskers show the 95% CI

**Supplementary Figure 2:**
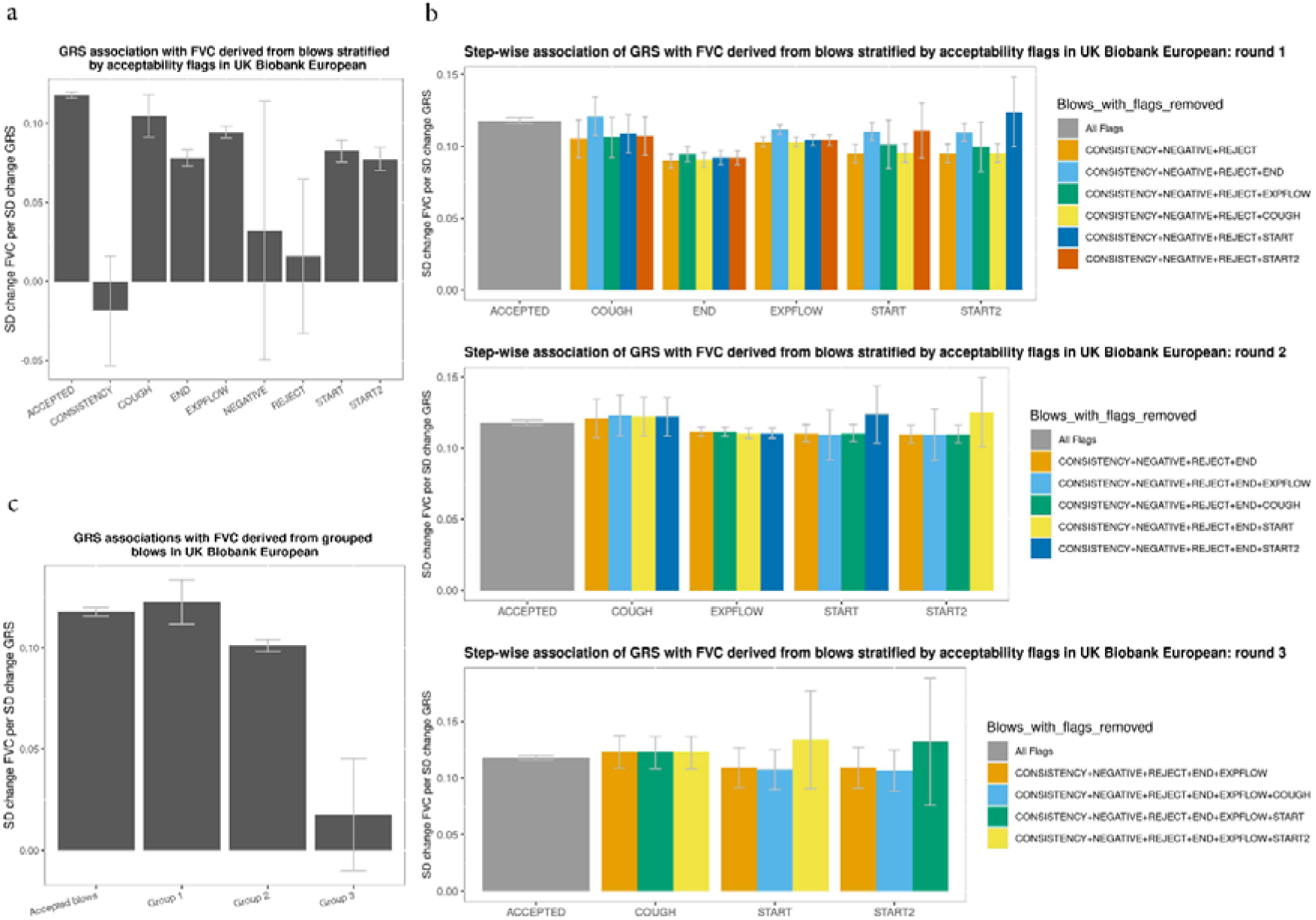
Ranking the impact of acceptability flags on spirometer blows. **a,** GRS association with FVC derived from spirometer blows stratified by acceptability flags shown as the s.d. change in FVC per s.d. increase in GRS. N.B. A blow could be included in multiple acceptability flag strata if it carries multiple acceptability flags. **b**, Iterative selection process. **c**, GRS association with FVC derived from grouped spirometer blows shown as the s.d. change in FVC per s.d. increase in GRS. Group 1 represents blows with hesitation flags only (“START” or “START2”). Group 2 represents blows with cough, end-of-blow or time to peak flow flags (“COUGH”, “END” or “EXPFLOW”) but without flags of “REJECT”, “CONSISTENCY” or “NEGATIVE”. Group 3 represents blows with acceptability flags of “REJECT”, “CONSISTENCY” or “NEGATIVE”. The height of the bars shows the point estimate of the effect and whiskers show the 95% CI

### Supplementary Tables

**Supplementary Table 1:** Sample size in 42 non-UKB cohorts.

| COHORT | ANCESTRY | N TOTAL | N MALES | N FEMALES |
| --- | --- | --- | --- | --- |
| ALHS | EUR | 2,848 | 1,447 | 1,401 |
| ALSPAC | EUR | 4,857 | 2,444 | 2,413 |
| ARIC-EA | EUR | 9,275 | 4,376 | 4,899 |
| B58C | EUR | 5,788 | 2,862 | 2,926 |
| BHS | EUR | 4,272 | 1,884 | 2,388 |
| CHS-EA | EUR | 3,220 | 1,259 | 1,961 |
| Croatia-Korcula | EUR | 2,520 | 929 | 1,591 |
| Croatia-Split | EUR | 961 | 379 | 582 |
| Croatia-Vis | EUR | 925 | 390 | 535 |
| EPIC-Norfolk | EUR | 20,771 | 9,664 | 11,107 |
| EXCEED | EUR | 1,334 | 818 | 516 |
| FinnTwin | EUR | 539 | 91 | 448 |
| FHS | EUR | 7,905 | 3,656 | 4,249 |
| GS | EUR | 16,048 | 6,633 | 10,415 |
| H2000 | EUR | 3,808 | 1,655 | 2,153 |
| HUNT | EUR | 16,485 | 7,648 | 8,837 |
| KORA-F4 | EUR | 1,474 | 717 | 757 |
| KORA-S3 | EUR | 1,147 | 551 | 596 |
| LBC1936 | EUR | 1,002 | 509 | 493 |
| MESA-CAU | EUR | 1,397 | 707 | 690 |
| NFBC1966 | EUR | 5,078 | 2,417 | 2,661 |
| NFBC1986 | EUR | 3,210 | 1,516 | 1,694 |
| SAPALDIA-Edinburgh | EUR | 2,584 | 1,338 | 1,246 |
| SAPALDIA-Gabriel | EUR | 1,355 | 656 | 699 |
| Orcades | EUR | 1,821 | 730 | 1,091 |
| PIVUS | EUR | 806 | 395 | 411 |
| Raine Study | EUR | 1,213 | 637 | 576 |
| RS1 | EUR | 1,240 | 526 | 714 |
| RS2 | EUR | 1,143 | 532 | 611 |
| RS3 | EUR | 1,431 | 624 | 807 |
| SHIP-START | EUR | 1,759 | 860 | 899 |
| SHIP-TREND | EUR | 2,519 | 1,265 | 1,254 |
| TwinsUK | EUR | 4,227 | 380 | 3,847 |
| UKHLS | EUR | 7,442 | 3,290 | 4,152 |
| VIKING | EUR | 1,701 | 672 | 1,029 |
| YFS | EUR | 419 | 198 | 221 |
| ARIC-AA | AFR | 2,820 | 1,044 | 1,776 |
| CHS-AA | AFR | 635 | 238 | 397 |
| MESA-AFR | AFR | 908 | 439 | 469 |
| HCHS-SOL | AMR | 10,965 | 4,558 | 6,407 |
| MESA-HIS | AMR | 905 | 427 | 478 |
| CKB | EAS | 83,715 | 36,399 | 47,316 |

**Supplementary Table 2:**
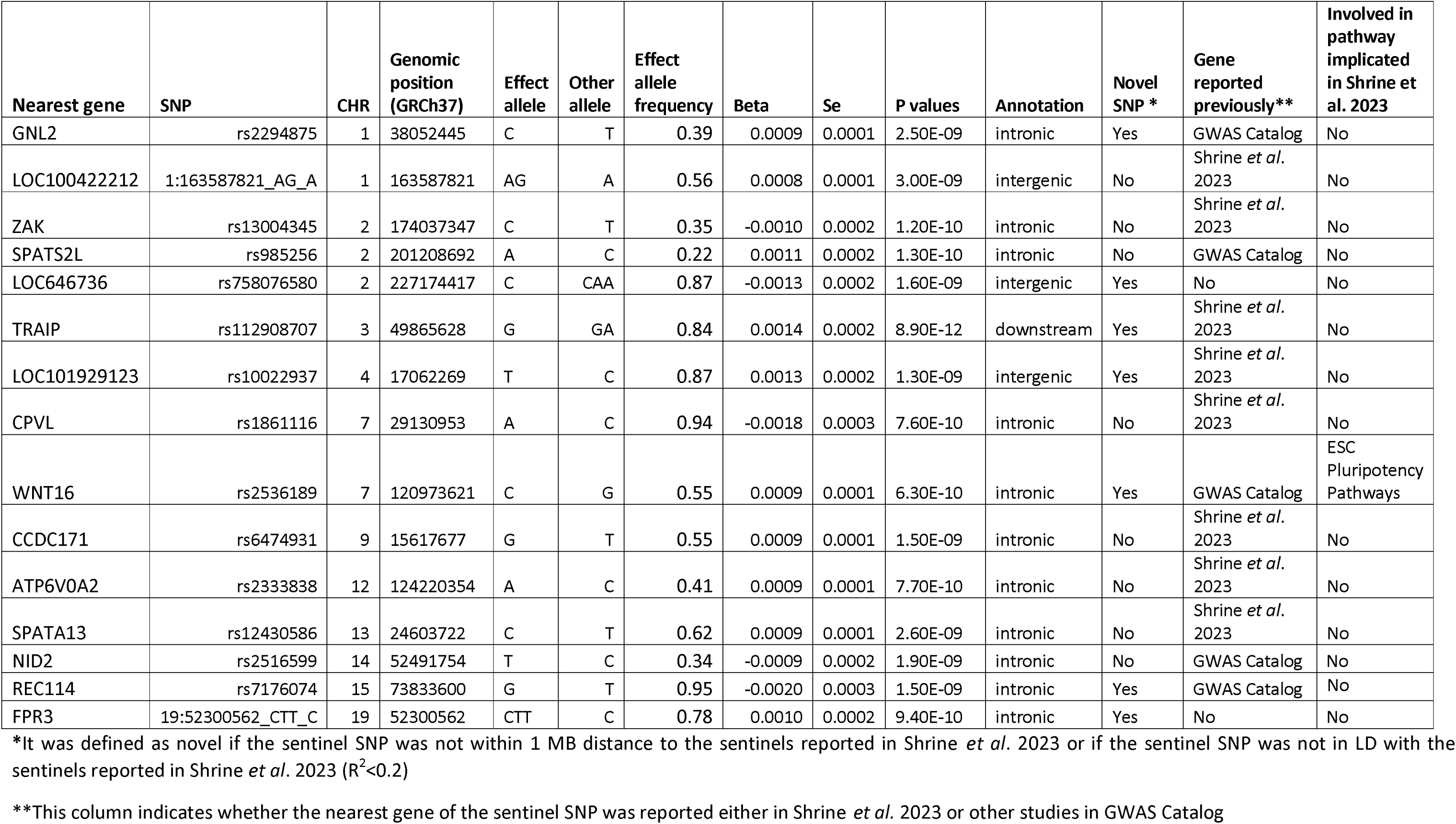
Additional signals for FEV_1_/FVC using relaxed spirometry QC criteria.

| Nearest gene | SNP | CHR | Genomic position (GRCh37) | Effect allele | Other allele | Effect allele frequency | Beta | Se | P values | Annotation | Novel SNP * | Gene reported previously** | Involved in pathway implicated in Shrine et al. 2023 |
| --- | --- | --- | --- | --- | --- | --- | --- | --- | --- | --- | --- | --- | --- |
| GNL2 | rs2294875 | 1 | 38052445 | C | T | 0.39 | 0.0009 | 0.0001 | 2.50E-09 | intronic | Yes | GWAS Catalog | No |
| LOC100422212 | 1:163587821_AG_A | 1 | 163587821 | AG | A | 0.56 | 0.0008 | 0.0001 | 3.00E-09 | intergenic | No | Shrine *et al.* 2023 | No |
| ZAK | rs13004345 | 2 | 174037347 | C | T | 0.35 | -0.0010 | 0.0002 | 1.20E-10 | intronic | No | Shrine *et al.* 2023 | No |
| SPATS2L | rs985256 | 2 | 201208692 | A | C | 0.22 | 0.0011 | 0.0002 | 1.30E-10 | intronic | No | GWAS Catalog | No |
| LOC646736 | rs758076580 | 2 | 227174417 | C | CAA | 0.87 | -0.0013 | 0.0002 | 1.60E-09 | intergenic | Yes | No | No |
| TRAIP | rs112908707 | 3 | 49865628 | G | GA | 0.84 | 0.0014 | 0.0002 | 8.90E-12 | downstream | Yes | Shrine *et al.* 2023 | No |
| LOC101929123 | rs10022937 | 4 | 17062269 | T | C | 0.87 | 0.0013 | 0.0002 | 1.30E-09 | intergenic | Yes | Shrine *et al.* 2023 | No |
| CPVL | rs1861116 | 7 | 29130953 | A | C | 0.94 | -0.0018 | 0.0003 | 7.60E-10 | intronic | No | Shrine *et al.* 2023 | No |
| WNT16 | rs2536189 | 7 | 120973621 | C | G | 0.55 | 0.0009 | 0.0001 | 6.30E-10 | intronic | Yes | GWAS Catalog | ESC Pluripotency Pathways |
| CCDC171 | rs6474931 | 9 | 15617677 | G | T | 0.55 | 0.0009 | 0.0001 | 1.50E-09 | intronic | No | Shrine *et al.* 2023 | No |
| ATP6V0A2 | rs2333838 | 12 | 124220354 | A | C | 0.41 | 0.0009 | 0.0001 | 7.70E-10 | intronic | No | Shrine *et al.* 2023 | No |
| SPATA13 | rs12430586 | 13 | 24603722 | C | T | 0.62 | 0.0009 | 0.0001 | 2.60E-09 | intronic | No | Shrine *et al.* 2023 | No |
| NID2 | rs2516599 | 14 | 52491754 | T | C | 0.34 | -0.0009 | 0.0002 | 1.90E-09 | intronic | No | GWAS Catalog | No |
| REC114 | rs7176074 | 15 | 73833600 | G | T | 0.95 | -0.0020 | 0.0003 | 1.50E-09 | intronic | Yes | GWAS Catalog | No |
| FPR3 | 19:52300562_CTT_C | 19 | 52300562 | CTT | C | 0.78 | 0.0010 | 0.0002 | 9.40E-10 | intronic | Yes | No | No |

\*It was defined as novel if the sentinel SNP was not within 1 MB distance to the sentinels reported in Shrine *et al.* 2023 or if the sentinel SNP was not in LD with the sentinels reported in Shrine *et al.* 2023 ( $R^2 < 0.2$ )
\*\*This column indicates whether the nearest gene of the sentinel SNP was reported either in Shrine *et al.* 2023 or other studies in GWAS Catalog

## Notes

### Competing Interest Statement

The authors have declared no competing interest.

### Funding Statement

Wellcome Trust Discovery Award (WT225221/Z/22/Z), NIHR Senior Investigator Award (NIHR201371) and the NIHR Leicester Biomedical Research Centre

### Author Declarations

UK Biobank has approval from the North West Multi-centre Research Ethics Committee (MREC) as a Research Tissue Bank (RTB) approval. This work was approved under project 648 in UK Biobank.

